# Effects of apolipoprotein B on the lifespan and risks of major disease including type 2 diabetes: a Mendelian randomization analysis using outcomes in first-degree relatives

**DOI:** 10.1101/2020.11.09.20227801

**Authors:** Tom G Richardson, Qin Wang, Eleanor Sanderson, Anubha Mahajan, Mark I McCarthy, Timothy M. Frayling, Mika Ala-Korpela, Allan Sniderman, George Davey Smith, Michael V Holmes

## Abstract

**Background:** Apolipoprotein B (apoB) is emerging as the lipoprotein entity that is critical for the role that lipoprotein lipids play in the aetiology of coronary heart disease (CHD). In this study, we explored effects of genetically-predicted apoB on endpoints in first-degree relatives.

**Methods:** Univariable Mendelian randomization (MR) used a weighted genetic instrument (229 SNPs) for apoB. For endpoints that apoB associated with at FDR <5%, multivariable MR analyses including genetic instruments for LDL-C and triglycerides. Estimates are inferred causal effects per 1-SD elevated lipoprotein trait (for apoB, 1-SD= 0.24 g/L). Replication of estimates for longevity and T2D was conducted using conventional two-sample MR using summary estimates from GWAS consortia.

**Findings:** Parents were less likely to be alive with 10.7 months of life lost in fathers (95%CI: 7.6, 13.9; FDR-adjusted P=4.0×10^−10^) and 5.8 months of life lost in mothers (95%CI: 3.0, 8.52; FDR-adjusted P=1.7×10^−4^) per 1-SD higher apoB in offspring. Effects strengthened to ∼2 yrs of life lost in multivariable MR and replicated in conventional two-sample MR (OR surviving to 90^th^ centile: 0.38; 95%CI: 0.22, 0.65). Genetically-elevated apoB caused higher risks of heart disease in all first-degree relatives and higher risk of stroke in mothers.

Findings in first-degree relatives were replicated in two-sample multivariable MR which identified apoB to increase (OR 2.32; 95%CI: 1.49, 3.61) and LDL-C lower (OR 0.34; 95%CI: 0.21, 0.54) risk of T2D.

**Interpretation:** Higher apoB shortens the lifespan, and increases risks of heart disease and stroke. T2D effects may represent injurious effects of dyslipidaemia to pancreatic islets.

**Research in Context:** *Evidence before this study:* Prior observational and Mendelian randomization studies have indicated that circulating concentrations of apoB are of critical importance to lipid-mediated atherogenesis, manifest as coronary heart disease.

*Added value of this study:* In this study, we explored the effects of genetically-predicted elevations in apoB on multiple endpoints occuring in first degree relatives including longevity and sought replication of findings using more conventional methods to exploit the statistical power from data available in large-scale GWAS consortia. We identified that apoB had a deleterious effect on longevity, shortening the lifespan by months to years. Furthermore, apoB caused higher risks of CHD and stroke in first degree relatives. Finally, apoB was identified to increase risk of T2D, in contradistinction to LDL-C which lowered risk of T2D, when employing multivairable MR methods.

*Implications of all the available evidence:* Our findings support apoB as being the major lipoprotein entity critical for CHD and stroke and extends this to identify higher apoB as negatively impacting longevity and increasing risk of T2D. These findings highlight the critical role of apoB in causing cardiometabolic disease, which collectively shortens the lifespan.

## Introduction

Blood lipid concentrations play an incontrovertible role in major vascular diseases, including coronary heart disease (CHD) and stroke ^1-5^. While LDL cholesterol is the conventional lipid trait of focus for observational and genetic epidemiological ^6,7^ studies and interventional trials ^8^, multiple sources of evidence support a central role of apolipoprotein B (apoB) in causing these effects ^9-12^.

The emergence of apoB as being of fundamental relevance in vascular disease has occurred on the background of decades of observational evidence including discordance analyses^13-15^, followed by genetic^16^ and Mendelian randomization ^17-19^ studies. These were facilitated by the development of statistical tools, including multivariable Mendelian randomization, which permits an appraisal of the causal effects of traits, while holding other traits in the same analysis constant, and without an introduction of collider bias ^20^. In brief, collider bias can arise when a trait is conditioned on (including the trait of being selected into a study), which can induce associations that undermine the validity of the analysis^21^.

While the role of apoB in coronary heart disease is increasingly emerging as of pivotal importance^9^, the comparative importance of apoB in other diseases remains less clear. This is important from an aetiological perspective, and also in light of drug development programmes where lipid-modifying therapeutic targets may have discrepant effects on lipoprotein particle constituents ^22,23^. In our previous analysis of the comparative role of atherogenic lipoprotein lipids, we focussed solely on CHD^18^. In this study, we expand the focus to explore a broader repertoire of outcomes, taking advantage of events reported as occuring in first-degree relatives in UK Biobank, thus facilitating investigations of multiple endpoints. The use of endpoints reported as occurring in first-degree relatives as the outcome in Mendelian randomization introduces several nuances into the interpretation of findings. For example, the genetic instrument is constructed in individuals who share ∼50% of the DNA to those in whom endpoints occur, which will lead to the expectation of a halving of the effect size for that which would be seen within individuals. However, such discovered associations can nonetheless provide insights into likely underlying causal relationships which, if consistent with conventional MR analyses (e.g. studies conducted within individuals, or through approximations to such using two-sample MR), bolster the confidence that the associations are real.

In this study, to appraise the role that apoB plays across major diseases, we explored the relationship of an apoB polygenic instrument with self-reported endpoints in first-degree relatives in participants of the UK Biobank. We assessed the rigor of these findings by conducting multivariable MR by including genetic instruments for other major atherogenic lipoprotein lipids (LDL-C and TG), and comparing findings to those previously published^17,18^. Finally, we sought replication of our findings with longevity and T2D in large-scale GWAS consortia.

## Methods

### Datasets and Ethics

Data on lipoprotein lipids and disease outcomes in first-degree relatives were obtained from the UK Biobank (UKBB) study under application #15825. Details on sample handling and assays for lipoprotein lipids in UKBB have been described previously^24^, as well as general characteristics of this cohort such as geographical regions and recruitment processes^25^. Likewise, details on genotyping quality control, phasing, imputation, and association testing in UKBB are reported elsewhere^26^. Ethical approval for this study was obtained from the Research Ethics Committee (REC; approval number: 11/NW/0382) and informed consent was collected from all participants enrolled in UKBB.

We sought replication of estimates from UKB using conventional two-sample MR approaches. Data for longevity were derived from a GWAS meta-analysis of cohorts with individuals dichotomised according to whether they survived to an age corresponding to the 90^th^ centile of longevity^27^. Data for T2D were derived from the most recent large-scale European T2D GWAS that included data from 32 studies and excluded participants from the UK Biobank study^28^.

### Derivation of genetic instruments for summary-level MR

Each lipoprotein lipid trait was normalized to have a mean of 0 and standard deviation of 1 using inverse rank-normalization. We undertook GWAS on all UKBB participants with genotype and trait data after excluding individuals of non-European descent (based on K-mean clustering of K=4) and standard exclusions, such as withdrawn consent, mismatch between genetic and reported sex, and putative sex chromosome aneuploidy. The BOLT-LMM (linear mixed model) software was selected to conduct GWAS due to its robustness to potential confounding due to relatedness or population structure^29,30^. Analyses were adjusted for age, sex, fasting status and a binary variable denoting the genotyping chip used in individuals (the UKBB Axiom array or the UK BiLEVE array).

Genetic instruments for each lipid-related trait were derived by undertaking linkage disequilibrium (LD) clumping of GWAS results. This involved identifying independent genetic variants robustly associated with traits (based on P<5×10^−8^) using a reference panel of 503 Europeans from phase 3 (version 5) of the 1,000 Genomes Project^31^. Clumping parameters of r2<0.001 and a distance of 1 MB were used for instrument identification. Univariable sets of instruments for each lipoprotein trait were derived by applying LD clumping on each trait’s GWAS results in turn. For multivariable instruments, we firstly combined all GWAS results together before applying LD clumping to ensure independent instruments were identified amongst these correlated traits.

### Outcomes

UKBB participants were asked at enrollment whether their father, mother or siblings suffered from any of the following diseases; heart disease, stroke, high blood pressure, chronic bronchitis/emphysema, Alzheimer’s disease/dementia, diabetes, Parkinson’s disease, severe depression, lung cancer, bowel cancer, prostate cancer or breast cancer. Additionally, individuals were asked whether their mother and father were still alive, and if not they were also asked at what age their parents had passed away. These questions were only eligible for participants who indicated that they were not adopted as a child and specifically regarded blood relations only. If there was any uncertainty over responses, participants were told to respond with ‘do not know’.

Each disease outcome for father, mother and sibling were analysed using GWAS separately based on BOLT-LMM and the QC pipeline described previously^32^. Total numbers of cases and controls for each of these GWAS can be found in **Table S1**. Genetic estimates for all instruments were extracted from these GWAS results for two-sample MR analyses. In other words, although UKB permits the application of Mendelian randomization using statistical approaches suited to the use of individual participant data, we in effect conducted analyses using a two-sample design (although the sample for the exposure and outcomes originated from the same dataset). Given that genetic variants for the instrument were identified from measurements in individuals separate to those that experienced disease (i.e. the SNP to exposure and SNP to outcome estimates were derived from different ‘people’), the potential for overfitting should theoretically be reduced. However, to address issues of potential overfitting leading to false positives, we sought to replicate key findings, as described below, using data on outcomes from non-overlapping datasets.

### Analyses

To quantify instrument strength, we calculated mean F-statistics to evaluate the instrument strengths in both the univariable and multivariable MR using the approximation described by Bowden et al^33^ and the conditional F-statistic by Sanderson et al ^34^ respectively.

We firstly investigated the effect of genetically predicted apoB on each of the 38 first-degree relative outcomes (described in **Table S1**) in turn using univariable MR. Genetic effects on apoB and outcomes for each instrument were harmonized and initial analyses were conducted using the inverse variance weight (IVW) method. Proxy SNPs were not necessary for this analysis given that both exposure and outcome datasets were from UKB. All univariable MR analyses were repeated for each outcome using LDL cholesterol and subsequently triglycerides as our exposure. Next, we conducted multivariable MR analyses to investigate the effects of apoB, LDL-C and triglycerides simultaneously on first-degree relative outcomes. This permitted us to estimate the ‘direct effect’ of apoB and allowed us to investigate the comparative causal roles of atherogenic lipoprotein traits on lifespan and disease risk, as proxied by outcome data on first-degree relatives.

Estimates from MR are presented as odds ratios per SD higher lipid trait. This also applies to effect estimates pertaining to the relationship of lipid traits with vital status and age at death of parents. This is because, by being obtained from UKB individuals at study entry, such data on parental vital status are for all intents and purposes, cross-sectional in nature.

### Sensitivity analyses

For effects in the univariable MR analysis that survived FDR<5% corrections based on estimates from the IVW method, we applied the weighted median^35^, weighted mode^36^ and MR-Egger^37^ regression approaches. For multivariable analyses^20^, we additionally conducted multivariable MR-Egger^38^.

To replicate the findings identified for longevity and T2D, we undertook a conventional 2-sample MR analysis in which SNPs for exposures (apoB, LDL-C and TG) were derived from UKB participants and SNPs for longevity (grouping study participants based on whether they survived to the age corresponding to the 90^th^ centile) and T2D were derived from recent large-scale GWAS^27,28^. The GWAS for longevity used UKB as a form of validation and the estimates we use here do not include UKB. However, the latest release of data from DIAMANTE included T2D cases from UKB (and including this might lead to over-fitting of the MR estimate), thus we analysed T2D GWAS estimates on individuals of European descent with UK Biobank removed. We conducted conventional univariable IVW 2-sample MR. We then fitted a multivariable MR model simultaneously including genetic instruments for apoB, LDL-C and TG to explore their independent causal roles. This was followed by a MVMR-Egger^38^ sensitivity analysis as a means of evaluating whether the MR estimates were influenced by dose-response confounding of the genetic instruments.

Because the apoB effects on risk of T2D might be confounded by adiposity and fat distribution acting through a residual VLDL element (not captured by conventional measurements of TG or LDL-C), we additionally included body mass index (BMI) and waist-hip-ratio adjusted for BMI (WHRadjBMI) respectively in the multivariable MR analyses for T2D. 641 SNPs used in the instrument for BMI were derived from n=461,377 UK Biobank participants using the same GWAS pipeline as described previously ^18^. 398 SNPs were used to instrument WHRadjBMI using previously undertaken GWAS within UK Biobank^39^. We also undertook analyses to evaluate the effect of T2D genetic liability on apoB levels using the IVW, MR-Egger and weighted median approaches in a univariable setting to investigate the potential bi-directional relationship between apoB and T2D.

### Multiple testing

Because first-degree relatives share ∼50% of their DNA, and because endpoints were reported multiple times among first degree relatives, the exposure to outcome analyses are not truly independent and use of conventional Bonferroni approaches to address multiple testing would therefore be overly stringent. We therefore used Benjamini-Hochberg FDR<5% to guide our interpretation of results that emerged from the initial univariable MR analysis of apoB onto all 38 outcomes. This heuristic was used to highlight outcomes to evaluate in further detail in all subsequent analyses. FDR corrections were also applied to all additional univariable and multivariable analyses using UKB participant data for completeness.

### Software

The BOLT-LMM software^29^ was used to undertake GWAS analyses and identify genetic instruments. We used the ‘TwoSampleMR’ R package to undertake all MR analyses^40^. The ‘ggplot2’ R package was used to generate forest plots^41^. Conditional F-statistics were generated using software from https://github.com/WSpiller/MVMR.

## Results

Up to 454,999 UK biobank participants reported information on prevalent diseases in first degree relatives. A median of 361,816 reported data for their 1^st^-degree siblings, with the corresponding values for mothers and fathers being 423,692 and 400,687, respectively. **Table S1** provides precise numbers of 1^st^-degree relatives for which endpoints were reported by UK biobank participants. 39.7% of UK biobank participants reported that their mothers were alive and 23.1% their fathers, with the mean age at death among those parents that had died being 75.7 yrs (SD 13.3) in mothers and 70.9 yrs (SD 13.1) in fathers.

Across the diseases reported as occurring in first-degree relatives (**Table 1**), the prevalence was similar in fathers (range 2.5-32.7%) and mothers (range 1.6-30.7%), with siblings having the lowest prevalence of disease (0.6-21.1%). Mothers were disproportionately affected by some diseases (e.g. 8.6% of mothers had Alzheimer’s disease/dementia as compared to 4.8% of fathers; 6.7% of mothers had severe depression compared to 3.9% of fathers). The prevalence of heart disease was highest in fathers (32.7%), with mothers (20.1%) and siblings (10.4%) being less affected. The prevalence of stroke was more similar among fathers (15.6%) and mothers (14.3%) but low in siblings (3.3%) whereas the prevalence of T2D was interestingly similar in all 1^st^-degree relatives (9.7% in fathers, 9.5% in mothers and 8.6% in siblings.).

**Table 1.** Vital status, age at death and prevalence of outcomes in first-degree relatives, as reported by UK Biobank participants. % cases unless otherwise stated; * age at death for fathers of 341,118 UKB participants and mothers of 273,111 UKB participants.. § value corresponds to median number of UKB participants reporting any endpoint in fathers, mothers or siblings (see **Table S1** for further details)

| **Outcome** | **Father**<br>(median<br>n=400,687§) | **Mother**<br>(median<br>n=423,692§) | **Siblings**<br>(median<br>n=361,816§) |
| --- | --- | --- | --- |
| Age at death, mean (s.d) * | 70.9 (13.1) | 75.7 (13.3) | N/A |
| Still alive | 23.08 | 39.66 | N/A |
| Alzheimer's disease/dementia | 4.82 | 8.63 | 0.58 |
| Bowel cancer | 5.97 | 5.21 | 2.47 |
| Breast cancer | N/A | 8.29 | 4.58 |
| Chronic bronchitis/emphysema | 11.50 | 5.97 | 2.85 |
| Diabetes | 9.70 | 9.46 | 8.56 |
| Heart disease | 32.71 | 20.09 | 10.41 |
| High blood pressure | 22.65 | 30.71 | 21.13 |
| Lung cancer | 9.32 | 4.15 | 2.27 |
| Parkinson's disease | 2.53 | 1.66 | 0.56 |
| Prostate cancer | 7.74 | N/A | 1.65 |
| Severe depression | 3.86 | 6.70 | 7.28 |
| Stroke | 15.60 | 14.33 | 3.32 |

### Genetic instruments

Our apoB GWAS in the UK Biobank explained 10.4% of the heritability in this trait. Undertaking LD clumping identified 229 apoB-associated SNPs with an F-statistic of 160. In the context of multivariable MR, where genetic instruments for apoB, LDL-C and TG were analysed simultaneously an additional 197 SNPs associated with LDL-C and 411 SNPS associated with TG were included in the analysis. Appraising the strength of the genetic instruments in multivariable MR, the conditional F-statistics for apoB and LDL-C were similar: for apoB the conditional F-statistic was 37, for LDL-C it was 34 and for TG it was 80. Thus in both univariable and multivariable MR settings, our genetic instruments would not be considered as weak.

### Relationship of apoB with participant-reported sibling and parental endpoints

Genetically-elevated apoB was associated with a lower probability that parents were alive (**Figure 1A** and **Table S2**). Fathers were likely to die 0.89 years (95%CI: 0.63, 1.16; FDR-adjusted P=4.0×10^−10^) earlier (corresponding to an average 10.7 months of life lost per 1-SD higher apoB in offspring) and mothers 0.48 years (95%CI 0.25, 0.71; FDR-adjusted P=1.7×10^−4^) earlier, corresponding to an average 5.8 months of life lost per 1-SD higher apoB in offspring. When taking into account the effects of LDL-C and TG in multivariable MR, these estimates for apoB strengthened to 1.94 years (95%CI: 0.91, 2.96; FDR-adjusted P=0.001) of life lost in fathers and 2.02 years (95%CI: 1.03, 3.01; FDR-adjusted P=3.9×10^−4^) of life lost in mothers, with the inverse relative odds for being alive also becoming more pronounced (**Figure 1B** and **Table S3**).

**Figure 1.**
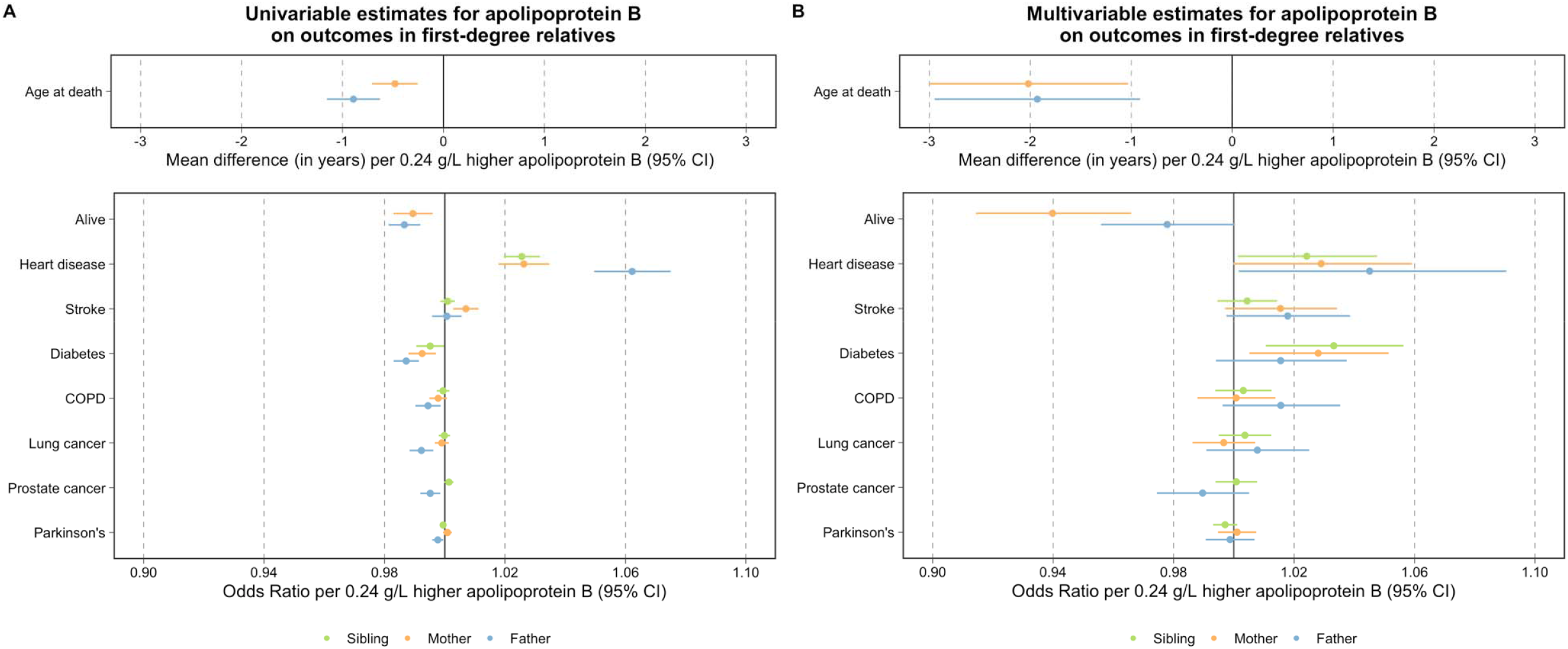
Univariable (A) and multivariable (B) Mendelian randomization estimates of genetically-elevated apolipoprotein B and risk of outcomes in first-degree relatives, including vital status and age at death. Univariable MR estimates displayed in (A). Multivariable MR estimates displayed in (B) represent the direct effects of apoB, adjusted for LDL-C and TG.

In univariable MR, a higher risk of heart disease was evident in all first-degree relatives of individuals with genetically-elevated apoB: OR in mother 1.03 (95%CI 1.02, 1.03; FDR-adjusted P=5.4×10^−9^), father 1.06 (95%CI: 1.05, 1.08; FDR-adjusted P=1.9×10^−21^) and siblings: 1.03 (95%CI: 1.02, 1.03; FDR-adjusted P=7.6×10^−16^). Mothers of individuals with genetically-elevated apoB had a higher risk of stroke (OR 1.007; 95%CI: 1.003, 1.011; FDR-adjusted P=4.2×10^−3^). Estimates remained similar, although less precise in multivariable MR (**Figure 1B**).

Parents of individuals with genetically elevated apoB had lower risks of T2D (OR in mothers 0.993; 95%CI 0.988; 0.997; FDR-adjusted P=0.004 and in fathers 0.987; 95%CI: 0.983, 0.991; FDR-adjusted P=2.3×10^−8^) with a trend to a commensurate lower risk in siblings (OR 0.995; 95%CI: 0.991, 0.9998; FDR-adjusted P=0.08). For all three estimates (i.e. the relationship of apoB with risk of T2D in mothers, fathers and siblings), the direction of effect was reversed – i.e. higher apoB was related to elevated T2D risk in multivariable MR: the relationship of the apoB genetic instrument with risk of T2D in mothers was OR 1.028 (95%CI: 1.005, 1.051; FDR-adjusted P=0.04) and in fathers it was OR 1.016 (95%CI: 0.994, 1.037; FDR-adjusted P=0.24).

### Univariable and multivariable MR estimates for LDL-C and TG

On univariable MR, the relationships of LDL-C with diseases in first-degree relatives were very similar to those of apoB for all endpoints (**Figure 2A** and **Table S4**). However, effect estimates changed on multivariable MR, with qualitative differences emerging in some instances: when taking into account apoB and TG in MVMR, higher LDL-C related to an older age at death in both parents (1.31 years in fathers; 95%CI: 0.22, 2.41 and 1.90 years in mothers; 95%CI: 0.84, 2.96; FDR-adjusted P=0.045 and 0.002, respectively) and a higher probability that mothers were alive (OR 1.06, 1.03, 1.09; FDR-adjusted P=7.6×10^−4^), with the estimate for fathers being weaker (OR 1.01; 95%CI: 0.99, 1.35; FDR-adjusted P=0.53) (**Figure 2B** and **Table S3**).

**Figure 2.**
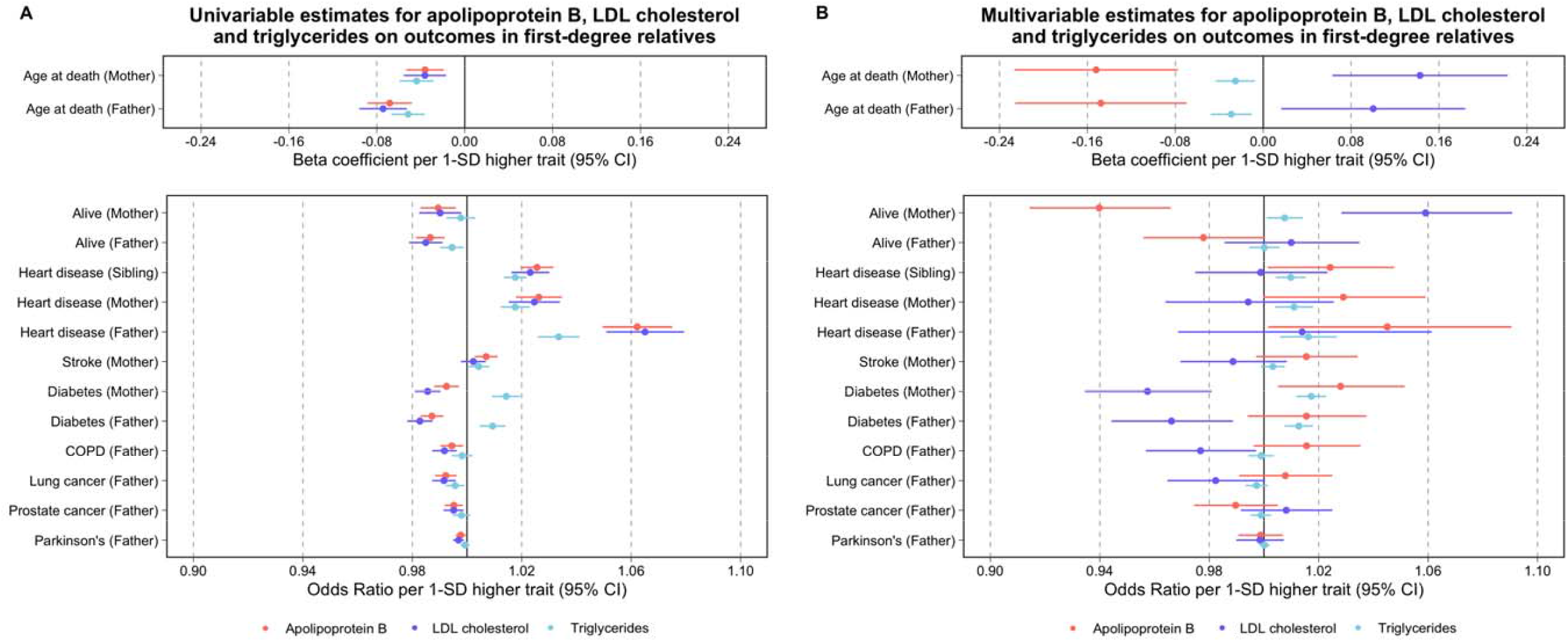
Univariable (A) and multivariable (B) Mendelian randomization estimates of genetically-elevated apolipoprotein B, LDL cholesterol and triglycerides and risk of outcomes in first-degree relatives, including vital status and age at death. Univariable MR estimates displayed in (A). Multivariable MR estimates displayed in (B) represent the direct effects of each lipoprotein entity, adjusted for the other two traits.

In contrast, the univariable and multivariable effects of LDL-C with risk of T2D remained directionally constant – i.e. higher LDL-C associated with a lower risk of T2D irrespective of whether genetic associations with apoB and TG were included in multivariable MR (fathers OR: 0.98; 95%CI: 0.98, 0.99 in univariable MR and OR 0.97; 95%CI: 0.94, 0.99 in MVMR; mothers OR: 0.99; 95%CI: 0.98, 0.99 in univariable MR and OR 0.96; 95%CI: 0.93, 0.98). The positive relationship of LDL-C with heart disease in parents and siblings (and with stroke in mothers) in univariable MR diminished markedly to the null on including apoB and TG, as did the inverse relationships with lung cancer, Parkinson’s and prostate cancer.

Estimates for TG were generally robust to multivariable MR analyses (**Figure 2, Table S5**). Higher TG related to an earlier age at death in both parents (0.68 fewer years in fathers; 95%CI: 0.47, 0.088; 0.58 fewer years lived in mothers; 95%CI: 0.38, 0.79; FDR-adjusted P=0.007 and 0.015, respectively) but the magnitude of association was comparatively smaller as compared to apoB in multivariable MR. Similarly, positive relationships between TG for heart disease in parents and siblings in univariable MR remained robust to multivariable MR but the magnitudes of effect were attenuated. For T2D, positive relationships of TG were evident in both univariable and multivariable MR analyses.

### Univariable and multivariable effects of apoB, LDL-C and TG on longevity and T2D using conventional two-sample MR

Given the findings we identified for vital status and risk of T2D in 1^st^-degree relatives, we sought to add additional information on these relationships under a conventional two-sample MR framework using data from two GWAS consortia: a GWAS of longevity with 11,262 individuals surviving to the age corresponding to the 90^th^ survival centile and DIAMANTE including 55,927 cases of T2D (**Figure 3**).

**Figure 3.**
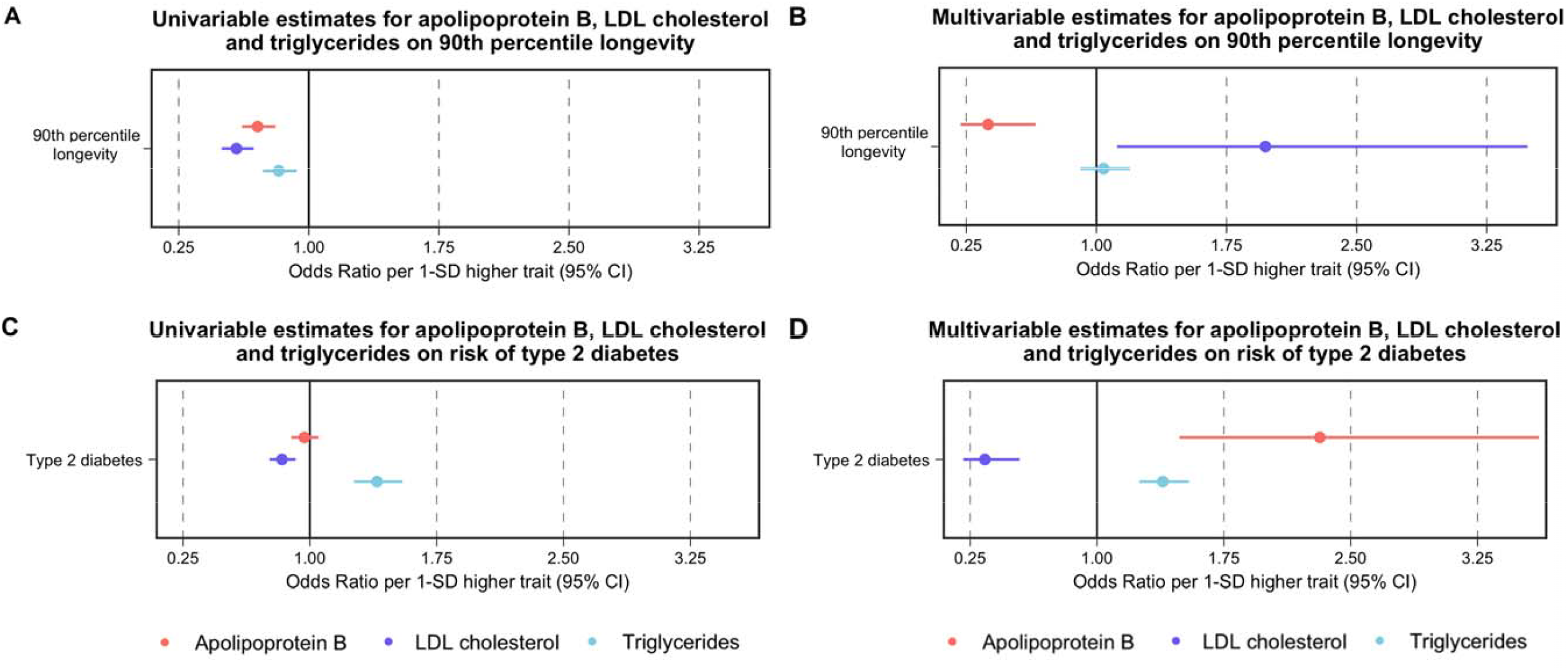
Univariable and multivariable estimates of genetically-elevated apolipoprotein B, LDL cholesterol and triglycerides with longevity (A and B) and risk of type 2 diabetes (C and D) using two-sample Mendelian randomization. Univariable MR estimates displayed in (A) and (C). Multivariable MR estimates displayed in (B) and (D) represent the direct effects of each lipoprotein entity, adjusted for the other two traits.

The analyses of longevity replicated findings from age of death and vital status of parents (**Figure 3** and **Table S12**). ApoB was strongly detrimental to survival (i.e. higher apoB caused individuals to die earlier than an age corresponding to the 90^th^ survival centile) and this relationship strengthened when taking into account the effects of LDL cholesterol and TG in multivariable MR. A 1-SD higher apoB in multivariable MR led to a 62% lower relative odds of surviving to the 90^th^ centile of longevity (corresponding to an OR of 0.38; 95%CI: 0.22, 0.65). In contrast, the initial harmful effect of LDL-C in univariable MR directionally reversed in multivariable MR. Likewise, TG had a detrimental effect on longevity on univariable MR but the relationship attenuated to the null on multivariable MR.

No clear effect of genetically-predicted apoB was identified for T2D in univariable MR. In contrast, higher LDL-C caused a lower risk of T2D (OR 0.84 per 1-SD higher LDL-C; 95%CI: 0.76, 0.92) and higher TG a higher T2D risk (OR 1.40 per 1-SD higher TG; 95%CI: 1.26, 1.55) in univariable MR. In multivariable MR, the estimate for TG remained largely unchanged (OR 1.39 per 1-SD higher TG; 95%CI: 1.25, 1.54). In strong contrast, the protective effect of LDL-C became more pronounced (OR 0.34; 95%CI: 0.21, 0.54) and a strong positive causal effect for apoB emerged (OR 2.32; 95%CI: 1.49, 3.61) (**Figure 3, Table S6**).

### Sensitivity analyses

Repeating analyses using robust MR approaches for both univariable and multivariable MR led to generally consistent relationships of apoB and risk of disease in first-degree relatives (**Figures S1-4** and **Tables S7-11**) and with risk of T2D in DIAMANTE (**Figure S5, Table S13**) and longevity (**Table S12**), The estimate of TG and risk of T2D was notable for being directionally opposite between IVW and MR-Egger in both univariable analyses for T2D in parents (**Figure S3**) and using conventional MR in DIAMANTE (**Figure S5, Table S13**). Of note, using MVMR-Egger and orientating SNPs so that they all associated with a higher TG led to an attenuation of the TG to T2D association suggesting that it might be explained by unbalanced horizontal pleiotropy (**Table S13**). Excluding variants from in/around the *APOE* locus led to a marked attenuation of the relationship between the apoB and LDL-C instrument and risk of Alzheimer’s disease in mothers (**Tables S14-S15**). Inclusion of body mass index in the MVMR for T2D in DIAMANTE had no major impact on the direct causal effects of apoB, LDL-C or TG (**Table S16** and **Figure S6)** which was also the case when accounting for WHRadjBMI **(Table S17** and **Figure S6**). We also applied univariable MR to evaluate evidence of a bi-directional relationship between apoB and T2D (i.e. whether T2D genetic liability influences apoB levels). We identified weak evidence of an effect in this analysis using genetic instruments derived from DIAMANTE based on the IVW and MR-Egger methods, whereas the weighted median estimate was directionally opposite to the MVMR estimate between apoB and T2D (**Table S18**).

## Discussion

In this study, we evaluated the relationship of genetically-elevated apoB with disease endpoints in first-degree relatives. The primary motivation was to explore the extent to which apoB related to major causes of disease and longevity. We further explored whether these relationships were resilient to including genetic instruments for the main atherogenic lipoprotein lipids, LDL cholesterol and triglycerides.

Our findings implicate apoB in several major diseases in first-degree relatives including heart disease, stroke and diabetes. Importantly, our findings show higher apoB to be linked to a shorter lifespan in parents as measured by both a lower relative odds that an individual’s mother and father are alive, and a younger age at their death – with up to 2 years of life lost per 1-SD higher apoB. Notably estimates tended to become more pronounced on inclusion of LDL cholesterol and triglycerides in multivariable MR, suggesting that once the effects of LDL-cholesterol and TG are taken into account, apoB was the predominant driving influence on lifespan among these three traits. These findings were replicated using an independent dataset further verifying the robustness of our findings. This provides a strong message that reductions in apoB should be the primary goal of lipid lowering as not only does this lead to lower risk from common diseases such as heart disease, stroke and T2D, but that the totality of effect is that, a reduction in apoB prolongs life by months to years. As discussed below, the effect estimates we report in this study, while approximating causal effects, are diluted owing to the nature of the exposure and outcome, which means that the real magnitudes of effect, in terms of duration of life lost due to elevated apoB, are likely to be considerably greater, as manifest in the effect estimates from the GWAS of 90^th^ centile of survival. These findings add important information to whether drugs that modify apoB are beneficial if the overarching aim is to lower risk of common disease and prolong life.

Our findings in first degree relatives for heart disease replicate those previously published using 2-sample MR using data from both the US National Center for Biotechnology Information Database of Genotypes and Phenotypes program^42^ (dbGAP)^17^ and UK biobank^43^ and demonstrate that in the presence of genetic variants associated with LDL-C and TG, apoB retains a positive effect on heart disease while the estimates for LDL-C are very much diminished. This congruence of findings further strengthens the evidence base that it is the number of circulating apoB particles, rather than the relative lipid content, that is the critical element for atherogenesis manifest as CHD and ischemic stroke. ^9,10,12,44^ As previously discussed^18^, this does not mean that LDL-C does not cause cardiovascular disease, but rather, frames the interpretation of lipid lowering therapy to focus on apoB as the driving force behind lipid-mediated atherosclerosis.

The findings for T2D are intriguing. On naive univariable MR analysis, a higher LDL-C was associated with a reduced risk of T2D, a finding, that strengthened with multivariable MR analysis. These results are consistent with studies employing MR using LDL-C genetic instruments^45^ that show higher LDL-C to be linked to lower risk of T2D. By contrast, apoB appeared to be associated with lower risk of T2D on univariable meta-analysis, but on multivariable MR, a strong, positive relation between apoB and risk of T2D emerged. This finding was replicated using data from DIAMANTE providing confirmatory evidence of a potential direct causal relation between apoB and T2D.

How might the contradictory trends between LDL-C, on the one hand, and apoB, on the other, to the risk of T2D be explained? LDL particles are the metabolic products of VLDL metabolism in the circulation and are primarily cleared by the LDL pathway. While the liver is, by far, the major site of clearance of LDL particles from plasma, peripheral tissues, such as the pancreas, do have LDL receptors and do clear LDL particles ^46^ (**Figure 4A**). Moreover, there is evidence that excess uptake of LDL particles injures islet cells ^47-52^ and adipocytes ^53,54^. What then are the determinants of LDL particle number in plasma and LDL clearance from plasma? LDL particle number in plasma may be increased, either because LDL clearance by the LDL pathway is reduced or because hepatic secretion of VLDL particles is increased, resulting in increased production of LDL particles ^55^. Decreased LDL clearance is characterized by LDL particles that are cholesterol enriched ^56,57^. In these circumstances, circulating LDL-C is disproportionately higher than apoB (**Figure 4B**): this is the scenario approximated by our multivariable MR analysis when LDL-C is increased and apoB kept constant. To the extent LDL clearance is reduced, uptake of LDL particles by pancreatic islet cells will be reduced and the potential for cell injury due to accumulation of cholesterol reduced. Familial hypercholesterolemia represents the extreme of decreased LDL clearance and, interestingly, patients with familial hypercholesterolemia have a lower incidence of T2D ^58^. Accordingly, reduced LDL clearance by pancreatic islet cells may account for the inverse relation between LDL-C and the risk of diabetes we observe.

**Figure 4.**
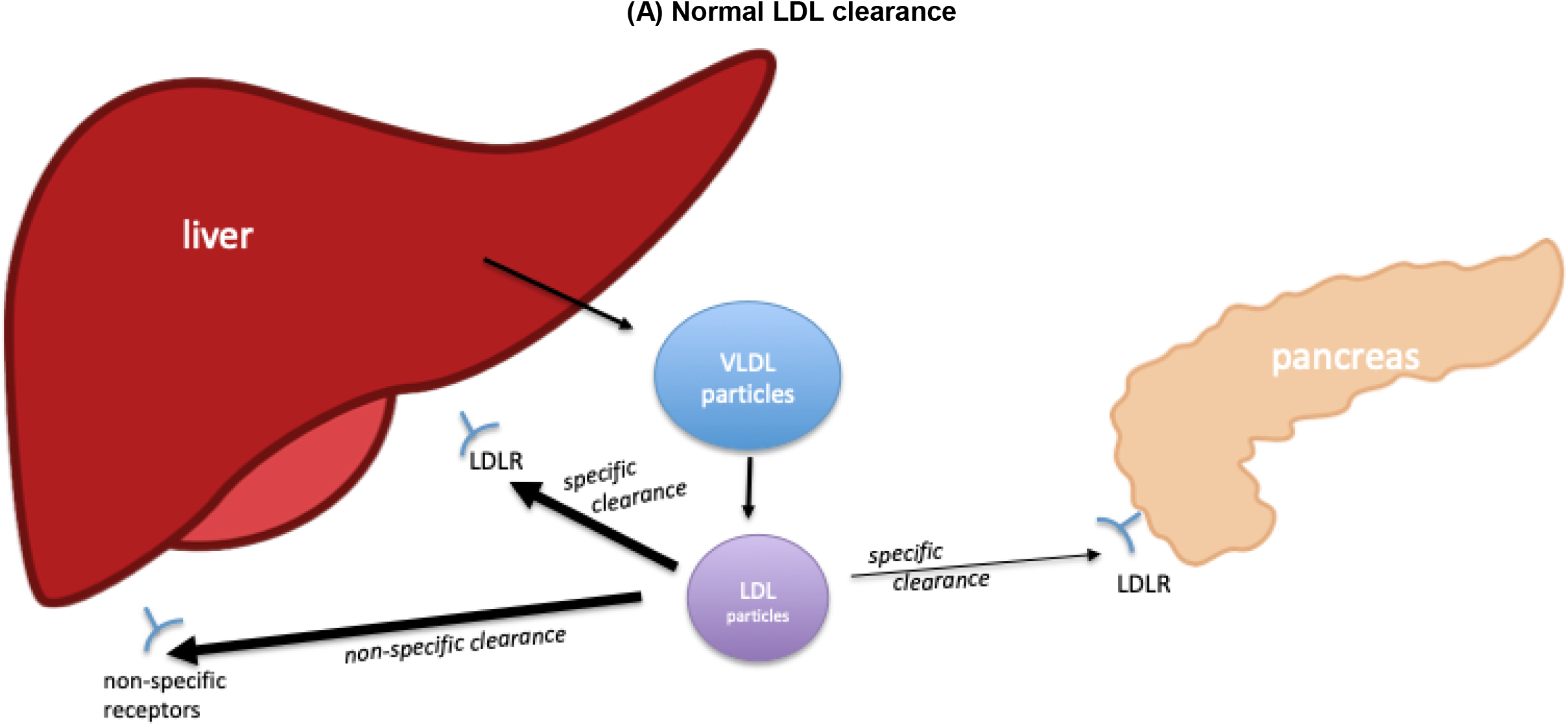
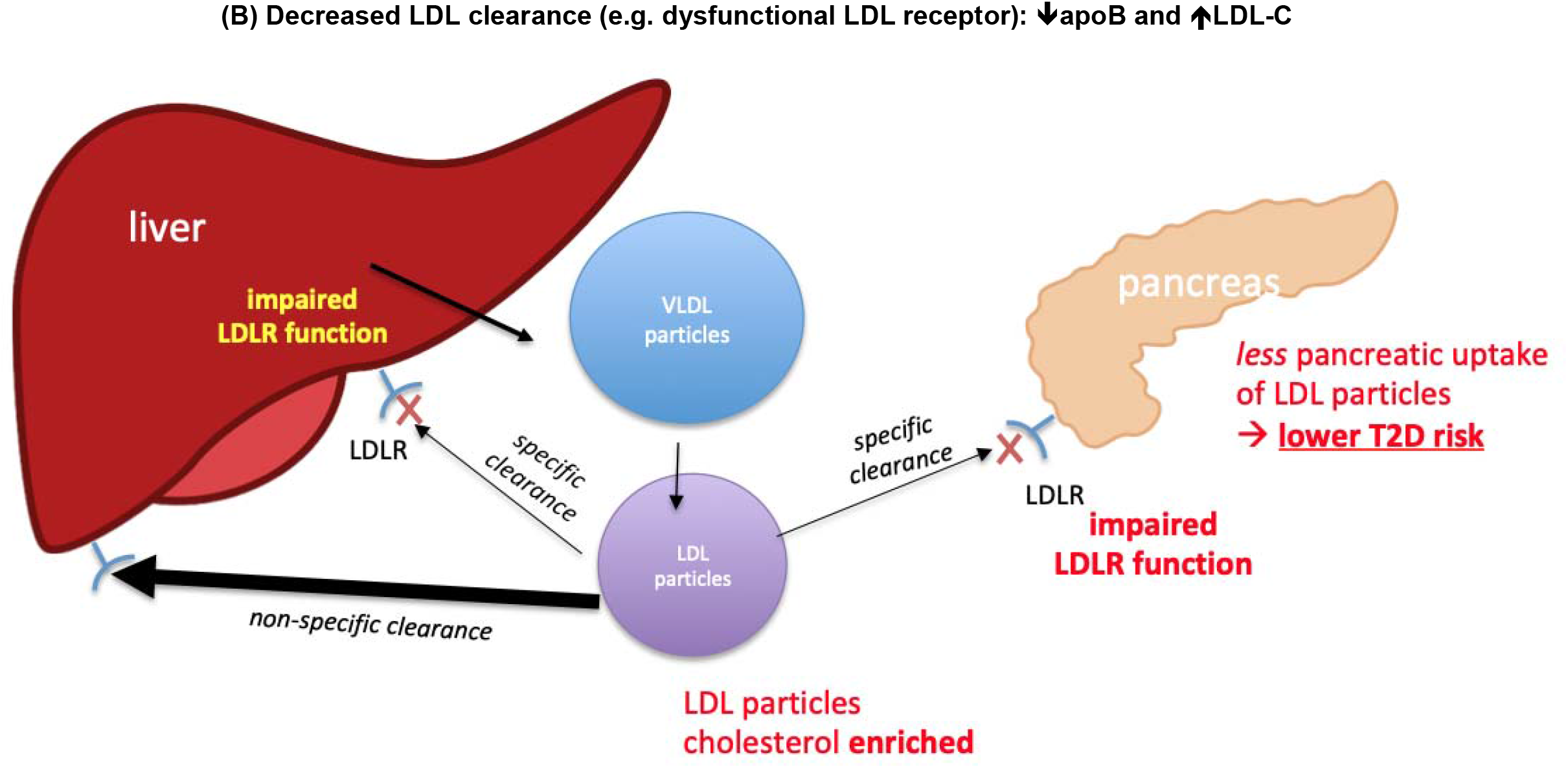
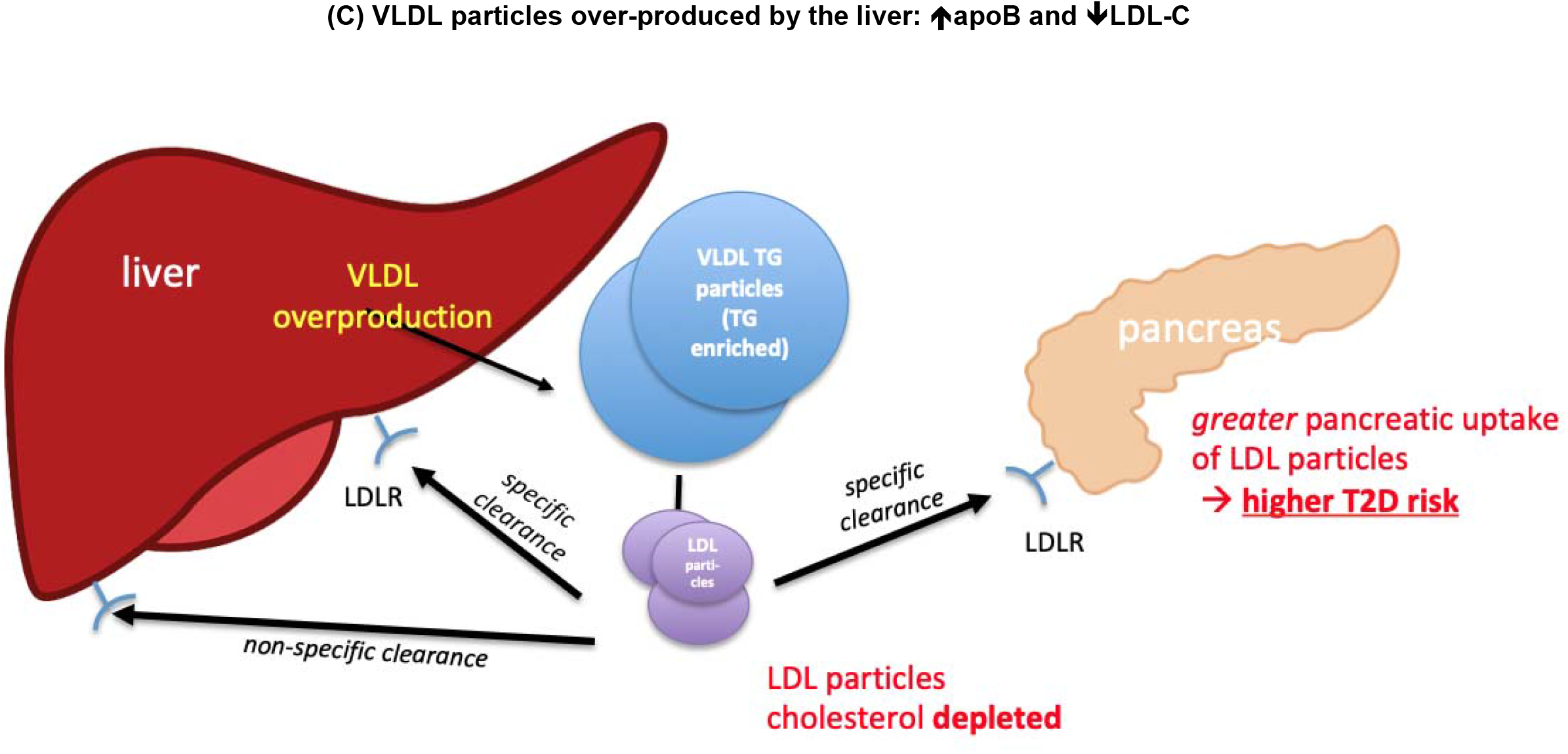
Metabolism of low density lipoprotein under (A) normal physiological conditions and abnormal physiological conditions of: (B) impaired LDL particle uptake by LDL receptor pathway, and; (C) over-production of VLDL particles. **Panel A: metabolism of low density lipoprotein (LDL) under normal physiological processes**. The liver synthesizes and secretes VLDL particles which, with removal of triglyceride within them, are transformed into LDL particles. LDL particles are cleared principally by the liver via specific LDL receptors (LDLR), which are concentration-dependent and saturable, and non-specific receptors, which are concentration-independent but non-saturable. As LDLR number decreases, a greater proportion of LDL clearance occurs via non-specific clearance. However, a small portion of LDL particles are cleared by LDLR in peripheral cells, such as the pancreas. Even though the uptake of LDL particles by the pancreas represents a trivial portion of total apoB clearance, we posit that variations in activity of this pathway may lead to altered risk of T2D. **Panel B**: impaired LDL receptor (LDLR) function leading to excess circulating LDL particles that are cholesterol enriched. Thus, apoB concentrations in plasma are comparatively lower than LDL-C – this scenario is approximated by our multivariable MR analysis when LDL-C is increased and apoB kept constant. In this circumstance, clearance of LDL from the circulation occurs principally through non-specific pathways. The reduction in LDL clearance through specific pathways (via LDLR) leads to a reduction in uptake of LDL particles by the pancreas. Because LDL uptake in excess can be injurious to beta cell function in the islets of Langerhans within the pancreas, reduced clearance leads to a lower risk of T2D. **Panel C**: VLDL over-production by the liver. This leads to higher circulating LDL particles that are comparatively cholesterol depleted. In other words, circulating apoB concentrations are comparatively higher than LDL-C – this scenario is approximated by our multivariable MR analysis when apoB is increased and LDL-C kept constant. In this scenario, overall LDL clearance is increased and occurs through both LDLR and non-specific pathways. Increased uptake of apoB particles by the pancreas leads to injury with commensurate reduction in function of beta cells and thus a higher risk of T2D.

By contrast, LDL particles tend to be depleted in cholesterol when LDL particles are overproduced, a common feature of hypertriglyceridemia, and a more common cause of increased LDL particle number than reduced LDL clearance. In such a circumstance, circulating apoB is disproportionately higher than LDL-C (**Figure 4C**), a scenario approximated by our multivariable MR analysis when apoB is increased and LDL-C kept constant and which points to increased concentrations of circulating cholesterol-depleted apoB-containing lipoprotein particles. In this instance, clearance of LDL particles by the LDL pathway is not impaired and indeed can be increased. Accordingly, delivery of LDL particles to pancreatic islet cells could be considerably increased. This pathophysiological sequence might explain, at least in part, the increased frequency of T2D and dysglycemia in these patients. Alternatively, increased uptake of LDL particles by adipocytes has been posited by Faraj^53^ to induce adipose tissue dysfunction with an increased inflammatory response resulting in reduced insulin sensitivity. Faraj^53^ and colleagues have also suggested cytotoxic injury to adipocytes due to increased uptake of LDL might be a mechanism by which statins and PCSK9 inhibitors, agents which increase activity of the LDL pathway, could be diabetogenic. Of note, PCSK9 deficiency in mice was associated with glucose intolerance due to reduced insulin secretion, and this was associated with larger islet cells in which there was accumulation of cholesterol esters.^47^ Accordingly, we hypothesize that increased uptake of LDL particles might injure adipocytes or islet cells or both and this might explain the powerful positive relation between apoB and the risk of diabetes that was observed in this study.

The resilience of our findings to inclusion of adiposity measured by BMI and WHRadjBMI (indexing total and visceral adiposity, respectively) in the multivariable MR suggests that the relationships we identify with risk of T2D are not confounded by upstream adiposity or fat distribution measures, which are recognized to causally influence blood lipid traits^59,60^. Thus, our findings may indicate new possibilities that therapies that lower pancreatic uptake of LDL particles while increasing hepatic uptake of LDL particles or precursor VLDL particles through other mechanisms might lead to a reduction in risk of both CHD and T2D.

Our study has several limitations. First, through the use of endpoints in first-degree relatives acquired through self-report of UK-biobank participants, we cannot claim that apoB directly causes these endpoints as the genetically-elevated apoB is only an approximation to unconfounded estimates *within* first-degree relatives. However, we can extrapolate that such estimates are likely a reflection of underlying causal structures, and the replication of our analyses for longevity and T2D corroborates our findings. Second, that 1st-degree relatives share 50% of their DNA means that by definition the genetic instruments we used have measurement error, leading to an attenuation of the effect estimates through regression dilution. This is evident, for example, by comparing the CHD, T2D and longevity associations using conventional 2-sample MR estimates as compared to estimates in first-degree relatives, where the magnitude is several-fold smaller, consistent with presence of regression dilution bias. Third, as endpoints are self-reported there is likely measurement error in such, and this may be differential by concurrent disease in UKB participants. For example, an individual who has heart disease may be more likely to report a 1^st^ degree relative as also having heart disease which would induce a relationship between the apoB genetic instrument (which causes CHD within the individual^18^) and endpoints in 1st-degree relatives. An additional feature is that although study participants were asked about blood relatives, they may have accidentally answered questions related to non-blood relatives (e.g. those adopted into families or step-parents/step-siblings) or half-siblings, which would have weakened effect estimates. Fourth, endpoints related to siblings may represent multiple individuals collapsed into a single trait (as individuals were only asked about siblings cumulatively rather than individually). This would dilute effects where more than one sibling has an endpoint (e.g. were two or more first-degree siblings to have CHD). The cumulative effect of these issues is likely to underlie why the effect estimates we report for heart disease (OR ∼1.03 to 1.05) are of considerably smaller magnitude than the approximation to “within-individual” effect estimates we previously reported using data from UKB and CARDIoGRAMplusC4D (OR ∼1.7) scaled to the same difference in exposure (∼1SD higher apoB in the UK biobank participant)^18^. A similar difference in the relative magnitudes of effect is evident between estimates of apoB with risk of T2D in 1^st^-degree relatives (OR ∼1.02) vs data from DIAMANTE (OR >2). Of note, both estimates for CAD (using CARDIoGRAMplusC4) and T2D (using DIAMANTE) may be inflated by spectrum-bias effects arising from case control studies that sample cases with ‘extreme phenotypes’ ^61^. Owing to the high genetic correlation between the traits explored (manifest in the comparatively lower conditional F-statistics for apoB and LDL-C as compared to triglycerides), the effect estimates derived from MVMR were imprecise. The nature of how vital status of parents was reported by UK biobank participants means that the variable is censored for parents who are alive (i.e. they are right censored^62^ – in other words, the event has yet to occur). This should be less of an issue in MR given that age causes censoring, and age was adjusted for in the GWAS. Finally, the protective effects of e.g. LDL-C with risk of T2D in parents could be reflective of a survival bias (i.e. individuals dying at a younger age from vascular disease, creating the impression of a lower risk of T2D that they would subsequently diagnose) ^63^. In this regard, findings from DIAMANTE (and the consistent association of apoB on MVMR in T2D reported in siblings) are reassuring as such estimates are less likely to be affected by survival bias.

The study also has several advantages. By using endpoints in first-degree relatives, we were able to investigate the relationship of apoB with endpoints such as vital status - while 6% of UKB participants have died during follow-up, 77% and 60% of UKB participant’s fathers and mothers had died. This permitted quantification of the relationships that we report for vital status and age at death. This study also provided a means to replicate prior work from us^18^ and others ^17^ showing the relative importance of apoB, LDL-C and TG to CHD. In addition, we were able to identify a T2D effect of apoB that directionally reversed when explored through MVMR, which has potential repercussions for development of therapies related to LDL particle clearance.

Another advantage here is that when effect estimates were consistent among first degree relatives, with previously published studies, and when using data from large-scale GWAS, they form a means of replication, strengthening the inference that can be made from the findings.

In conclusion, our evaluation of apoB using endpoints in first-degree relatives finds higher apoB to be detrimental to cardiovascular disease, diabetes and longevity. Lifestyle and pharmacological approaches to lowering apoB should have widespread beneficial effects, including preventing common diseases and prolonging life.

## Data Availability

Data will become available on publication (supplementary materials)

## Acknowledgements

We are immensely grateful to the study participants of the UKBB. TGR is a UKRI Innovation Research Fellow (MR/S003886/1). QW is supported by a postdoctoral fellowship from Novo Nordisk Foundation (NNF17OC0027034)’TMF has received funding from the Medical Research Council, MR/T002239/1 and the Innovative Medicines Initiative 2 Joint Undertaking under grant agreement No 875534. This Joint Undertaking support from the European Union’s Horizon 2020 research and innovation programme and EFPIA and T1D Exchange, JDRF, and Obesity Action Coalition. MMcC was a Wellcome Investigator supported by Wellcome funding (098381, 106130, 203141, 212259). MAK is supported by a research grant from the Sigrid Juselius Foundation, Finland. MVH works in a unit that receives funding from the UK Medical Research Council and is supported by a British Heart Foundation Intermediate Clinical Research Fellowship (FS/18/23/33512) and the National Institute for Health Research Oxford Biomedical Research Centre. GDS and TGR work within the Medical Research Council Integrative Epidemiology Unit at the University of Bristol MC_UU_00011/1.

## References

1. Boren J, Chapman MJ, Krauss RM, et al. Low-density lipoproteins cause atherosclerotic cardiovascular disease: pathophysiological, genetic, and therapeutic insights: a consensus statement from the European Atherosclerosis Society Consensus Panel. Eur Heart J 2020.

2. Collins R, Reith C, Emberson J, et al. Interpretation of the evidence for the efficacy and safety of statin therapy. The Lancet 2016; 388(10059): 2532–61.

3. Cholesterol Treatment Trialist’s Collaborators, Fulcher J, O’Connell R, et al. Efficacy and safety of LDL-lowering therapy among men and women: meta-analysis of individual data from 174,000 participants in 27 randomised trials. Lancet 2015; 385(9976): 1397–405.

4. Cholesterol Treatment Trialists’ Collaborators, Baigent C, Blackwell L, et al. Efficacy and safety of more intensive lowering of LDL cholesterol: a meta-analysis of data from 170,000 participants in 26 randomised trials. Lancet 2010; 376(9753): 1670–81.

5. Cholesterol Treatment Trialists’ Collaborators, Mihaylova B, Emberson J, et al. The effects of lowering LDL cholesterol with statin therapy in people at low risk of vascular disease: meta-analysis of individual data from 27 randomised trials. Lancet 2012; 380(9841): 581–90.

6. Ference BA, Majeed, F., Penumetcha, R., Flack JM, Brook RD. Effect of Naturally Random Allocation to Lower Low-Density Lipoprotein Cholesterol on the Risk of Coronary Heart Disease Mediated by Polymorphisms in NPC1L1, HMGCR, or Both. Journal of the American College of Cardiology 2015; 65(15): 1552–61.

7. Holmes MV, Asselbergs FW, Palmer TM, et al. Mendelian randomization of blood lipids for coronary heart disease. European Heart Journal 2015; 36(9): 539–50.

8. Silverman MG, Ference BA, Im K, et al. Association between lowering ldl-c and cardiovascular risk reduction among different therapeutic interventions: A systematic review and meta-analysis. JAMA 2016; 316(12): 1289–97.

9. Sniderman AD, Pencina M, Thanassoulis G. ApoB. Circ Res 2019; 124(10): 1425–7.

10. Sniderman AD, Thanassoulis G, Glavinovic T, et al. Apolipoprotein B Particles and Cardiovascular Disease: A Narrative Review. JAMA Cardiol 2019.

11. Sniderman AD, Williams K, Contois JH, et al. A meta-analysis of low-density lipoprotein cholesterol, non-high-density lipoprotein cholesterol, and apolipoprotein B as markers of cardiovascular risk. Circ Cardiovasc Qual Outcomes 2011; 4(3): 337–45.

12. Boren J, Williams KJ. The central role of arterial retention of cholesterol-rich apolipoprotein-B-containing lipoproteins in the pathogenesis of atherosclerosis: a triumph of simplicity. Curr Opin Lipidol 2016; 27(5): 473–83.

13. Wilkins JT, Li RC, Sniderman A, Chan C, Lloyd-Jones DM. Discordance Between Apolipoprotein B and LDL-Cholesterol in Young Adults Predicts Coronary Artery Calcification: The CARDIA Study. J Am Coll Cardiol 2016; 67(2): 193–201.

14. Sniderman AD, Islam S, Yusuf S, McQueen MJ. Discordance analysis of apolipoprotein B and non-high density lipoprotein cholesterol as markers of cardiovascular risk in the INTERHEART study. Atherosclerosis 2012; 225(2): 444–9.

15. Sniderman AD, Robinson JG. ApoB in clinical care: Pro and Con. Atherosclerosis 2019; 282: 169–75.

16. Ference BA, Kastelein JJP, Ginsberg HN, et al. Association of Genetic Variants Related to CETP Inhibitors and Statins With Lipoprotein Levels and Cardiovascular Risk. JAMA 2017; 318(10): 947–56.

17. Ference BA, Kastelein JJP, Ray KK, et al. Association of Triglyceride-Lowering LPL Variants and LDL-C-Lowering LDLR Variants With Risk of Coronary Heart Disease. JAMA 2019; 321(4): 364–73.

18. Richardson TG, Sanderson E, Palmer TM, et al. Evaluating the relationship between circulating lipoprotein lipids and apolipoproteins with risk of coronary heart disease: A multivariable Mendelian randomisation analysis. PLoS Med 2020; 17(3): e1003062.

19. Zuber V, Gill D, Ala-Korpela M, et al. High-throughput multivariable Mendelian randomization analysis prioritizes apolipoprotein B as key lipid risk factor for coronary artery disease. International Journal of Epidemiology 2020.

20. Sanderson E, Davey Smith G, Windmeijer F, Bowden J. An examination of multivariable Mendelian randomization in the single sample and two-sample summary data settings. Int J Epidemiol 2019; 48(3).

21. Munafò MR, Tilling K, Taylor AE, Evans DM, Davey Smith G. Collider scope: when selection bias can substantially influence observed associations. International Journal of Epidemiology 2017; 47(1): 226–35.

22. Holmes MV, Ala-Korpela M. What is ‘LDL cholesterol’? Nat Rev Cardiol 2019; 16(4): 197–8.

23. Holmes MV, Richardson TG, Ference BA, Davies N, Davey Smith G. Integrating genomics with biomarkers and therapeutic targets to invigorate cardiovascular drug development. Nature Reviews Cardiology 2020 (In Press).

24. Elliott P, Peakman TC, Biobank UK. The UK Biobank sample handling and storage protocol for the collection, processing and archiving of human blood and urine. Int J Epidemiol 2008; 37(2): 234–44.

25. Sudlow C, Gallacher J, Allen N, et al. UK biobank: an open access resource for identifying the causes of a wide range of complex diseases of middle and old age. PLoS Med 2015; 12(3): e1001779.

26. Bycroft C, Freeman C, Petkova D, et al. The UK Biobank resource with deep phenotyping and genomic data. Nature 2018; 562(7726): 203–9.

27. Deelen J, Evans DS, Arking DE, et al. A meta-analysis of genome-wide association studies identifies multiple longevity genes. Nat Commun 2019; 10(1): 3669.

28. Mahajan A, Taliun D, Thurner M, et al. Fine-mapping type 2 diabetes loci to single-variant resolution using high-density imputation and islet-specific epigenome maps. Nat Genet 2018; 50(11): 1505–13.

29. Loh PR, Tucker G, Bulik-Sullivan BK, et al. Efficient Bayesian mixed-model analysis increases association power in large cohorts. Nat Genet 2015; 47(3): 284–90.

30. Loh PR, Kichaev G, Gazal S, Schoech AP, Price AL. Mixed-model association for biobank-scale datasets. Nat Genet 2018; 50(7): 906–8.

31. Genomes Project C, Abecasis GR, Auton A, et al. An integrated map of genetic variation from 1,092 human genomes. Nature 2012; 491(7422): 56–65.

32. Mitchell R, Elsworth BL, Raistrick CA, Paternoster L, Hemani G, Gaunt TR. MRC IEU UK Biobank GWAS pipeline version 2. University of Bristol Available at: https://databrisacuk/data/dataset/pnoat8cxo0u52p6ynfaekeigi.

33. Bowden J, Del Greco MF, Minelli C, Davey Smith G, Sheehan NA, Thompson JR. Assessing the suitability of summary data for two-sample Mendelian randomization analyses using MR-Egger regression: the role of the I2 statistic. Int J Epidemiol 2016.

34. Sanderson E, Spiller W, Bowden J. Testing and Correcting for Weak and Pleiotropic Instruments in Two-Sample Multivariable Mendelian Randomisation. bioRxiv 2020.

35. Bowden J, Davey Smith, G., Haycock, P. C., Burgess, S. Consistent Estimation in Mendelian Randomization with Some Invalid Instruments Using a Weighted Median Estimator. Genetic Epidemiology 2016; 40(4): 304–14.

36. Hartwig FP, Davey Smith G, Bowden J. Robust inference in summary data Mendelian randomization via the zero modal pleiotropy assumption. Int J Epidemiol 2017; 46(6): 1985–98.

37. Bowden J, Davey Smith G, Burgess S. Mendelian randomization with invalid instruments: effect estimation and bias detection through Egger regression. Int J Epidemiol 2015; 44(2): 512–25.

38. Rees JMB, Wood AM, Burgess S. Extending the MR-Egger method for multivariable Mendelian randomization to correct for both measured and unmeasured pleiotropy. Stat Med 2017; 36(29): 4705–18.

39. O’Connor LJ, Price AL. Distinguishing genetic correlation from causation across 52 diseases and complex traits. Nature Genetics 2018; 50(12): 1728–34.

40. Hemani G, Zheng J, Elsworth B, et al. The MR-Base platform supports systematic causal inference across the human phenome. Elife 2018; 7.

41. Wickham H. ggplot2 – Elegant Graphics for Data Analysis (2nd Edition). Journal of Statistical Software 2017; 77.

42. Mailman MD, Feolo M, Jin Y, et al. The NCBI dbGaP database of genotypes and phenotypes. Nat Genet 2007; 39(10): 1181–6.

43. Richardson TG, Sanderson E, Elsworth B, Tilling K, Davey Smith G. Use of genetic variation to separate the effects of early and later life adiposity on disease risk: mendelian randomisation study. BMJ 2020; 369: m1203.

44. Ala-Korpela M. The culprit is the carrier, not the loads: cholesterol, triglycerides and apolipoprotein B in atherosclerosis and coronary heart disease. Int J Epidemiol 2019.

45. White J, Swerdlow DI, Preiss D, et al. Association of Lipid Fractions With Risks for Coronary Artery Disease and Diabetes. JAMA Cardiol 2016; 1(6): 692–9.

46. Dietschy JM, Spady DK, Stange EF. Quantitative importance of different organs for cholesterol synthesis and low-density-lipoprotein degradation. Biochem Soc Trans 1983; 11(6): 639–41.

47. Da Dalt L, Ruscica M, Bonacina F, et al. PCSK9 deficiency reduces insulin secretion and promotes glucose intolerance: the role of the low-density lipoprotein receptor. Eur Heart J 2019; 40(4): 357–68.

48. Cnop M, Hannaert JC, Grupping AY, Pipeleers DG. Low density lipoprotein can cause death of islet beta-cells by its cellular uptake and oxidative modification. Endocrinology 2002; 143(9): 3449–53.

49. Roehrich ME, Mooser V, Lenain V, et al. Insulin-secreting beta-cell dysfunction induced by human lipoproteins. The Journal of biological chemistry 2003; 278(20): 18368–75.

50. Hao M, Head WS, Gunawardana SC, Hasty AH, Piston DW. Direct effect of cholesterol on insulin secretion: a novel mechanism for pancreatic beta-cell dysfunction. Diabetes 2007; 56(9): 2328–38.

51. Kruit JK, Kremer PH, Dai L, et al. Cholesterol efflux via ATP-binding cassette transporter A1 (ABCA1) and cholesterol uptake via the LDL receptor influences cholesterol-induced impairment of beta cell function in mice. Diabetologia 2010; 53(6): 1110–9.

52. Ishikawa M, Iwasaki Y, Yatoh S, et al. Cholesterol accumulation and diabetes in pancreatic beta-cell-specific SREBP-2 transgenic mice: a new model for lipotoxicity. J Lipid Res 2008; 49(12): 2524–34.

53. Faraj M. LDL, LDL receptors, and PCSK9 as modulators of the risk for type 2 diabetes: a focus on white adipose tissue. J Biomed Res 2020; 34(4): 251–9.

54. Cyr Y, Bissonnette S, Lamantia V, et al. White Adipose Tissue Surface Expression of LDLR and CD36 is Associated with Risk Factors for Type 2 Diabetes in Adults with Obesity. Obesity (Silver Spring, Md) 2020.

55. Sniderman AD, De Graaf J, Couture P, Williams K, Kiss RS, Watts GF. Regulation of plasma LDL: the apoB paradigm. Clin Sci (Lond) 2009; 118(5): 333–9.

56. Teng B, Thompson GR, Sniderman AD, Forte TM, Krauss RM, Kwiterovich PO, Jr. Composition and distribution of low density lipoprotein fractions in hyperapobetalipoproteinemia, normolipidemia, and familial hypercholesterolemia. Proc Natl Acad Sci U S A 1983; 80(21): 6662–6.

57. Teng B, Sniderman AD, Soutar AK, Thompson GR. Metabolic basis of hyperapobetalipoproteinemia. Turnover of apolipoprotein B in low density lipoprotein and its precursors and subfractions compared with normal and familial hypercholesterolemia. J Clin Invest 1986; 77(3): 663–72.

58. Besseling J, Kastelein JJ, Defesche JC, Hutten BA, Hovingh GK. Association between familial hypercholesterolemia and prevalence of type 2 diabetes mellitus. JAMA 2015; 313(10): 1029–36.

59. Holmes MV, Lange LA, Palmer T, et al. Causal effects of body mass index on cardiometabolic traits and events: a Mendelian randomization analysis. Am J Hum Genet 2014; 94(2): 198–208.

60. Dale CE, Fatemifar G, Palmer TM, et al. Causal Associations of Adiposity and Body Fat Distribution With Coronary Heart Disease, Stroke Subtypes, and Type 2 Diabetes Mellitus: A Mendelian Randomization Analysis. Circulation 2017; 135(24): 2373–88.

61. Gan W, Walters RG, Holmes MV, et al. Evaluation of type 2 diabetes genetic risk variants in Chinese adults: findings from 93,000 individuals from the China Kadoorie Biobank. Diabetologia 2016; 59(7): 1446–57.

62. Sedgwick P. Survival (time to event) data: censored observations. BMJ 2011; 343.

63. Mitchell BD, Kammerer CM, Reinhart LJ, Stern MP, MacCluer JW. Is there an excess in maternal transmission of NIDDM? Diabetologia 1995; 38(3): 314–7.

